# Modeling and Forecasting Trend of COVID-19 Epidemic in Iran until May 13, 2020

**DOI:** 10.1101/2020.03.17.20037671

**Authors:** Ali Ahmadi, Yasin Fadaei, Majid Shirani, Fereydoon Rahmani

## Abstract

**Background:** COVID-19 is an emerging disease and precise data are not available in the world and Iran. this study aimed to determine the epidemic trend and prediction of COVID-19 in Iran.

**Methods:** This study is a secondary data analysis and modeling. We used the daily reports of definitive COVID-19 patients (sampling of severe cases and hospitalization) released by Iran Ministry of Health and Medical Education. Epidemic projection models of Gompertz, Von Bertalanffy and least squared error (LSE) were used to predict the number of cases at April 3, 2020 until May 13, 2020.

**Results:** R_0_ in Iran was estimated to be 4.7 that has now fallen to below 2. Given the three different scenarios, the prediction of the patients on April 3, 2020 by Von Bertalanffy, Gompertz and LSE were estimated at 48200, 52500 and 58000, respectively. The number of deceased COVID-19 patients was also estimated to be 3600 individuals using the Von growth model, 4200 ones by Gompertz’s model and 4850 ones according to the LSE method. To predict and estimate the number of patients and deaths in the end of epidemic based on Von and Gompertz models, we will have 87000 cases, 4900 and 11000 deaths until 13 May and 1 June, respectively.

**Conclusion:** The process of controlling the epidemic is tangible. If enforcement and public behavior interventions continue with current trends, the control and reduction of the COVID-19 epidemic in Iran will be flat from April 28, until July, 2020 and new cases are expected to decline from the following Iranian new year.

↑What is “already known” in this topic
COVID-19 is an emerging disease, pandemic and precise data on its epidemic are not available in the world and Iran.

→What this article adds
this study is shown, If enforcement and public behavior interventions continue with current trends, the control and reduction of the COVID-19 epidemic in Iran will be flat from April 28, until July, 2020.

## Introduction

Coronaviruses are a large family of viruses that have been identified since 1965 and so far 7 species of them have been discovered and reported to affect humans. These viruses have three genotypes of alpha, beta and gamma. The natural reservoirs of these diseases are mammals and birds, and therefore are considered as zoonotic diseases (1,2). Severe acute respiratory syndrome (SARS) is caused by a species of coronavirus that infects humans, bats and certain other mammals, which has led to epidemics in 2002 and 2003 (2-4), Middle East Respiratory Syndrome (MERS) caused 2012 epidemic in Saudi Arabia (5), and more recently their newest variant COVID-19 has led to the recent epidemic in China, Italia and then across the world (6-8). COVID-19 is an emerging disease, these viruses cause direct and indirect transmission of respiratory diseases with a wide range of asymptomatic, cold symptoms from respiratory/fever symptoms, cough, shortness of breath to kidney failure and death, leading to more severe respiratory illness epidemic and pandemic in most countries of the world (9). The complete clinical manifestation is not clear yet and understanding of the transmission risk is incomplete (10), but the virus is believed transmitted mostly via contact, droplets, aspirates, feces, and aerosols transmission is highly possible. individuals are all generally susceptible to the virus. the mean incubation period was 5.2 days (4.1–7 days), and the basic reproductive number (R_0_) was reported 2.2 (95% CI: 1.4 to 3.9) (11). In another study, the mean incubation period of COVID-19 was ranges from 0-24 days with mean of 6.4 days. The R_0_ of COVID-19 at the early phase regardless of different prediction models, which is higher than SARS and MERS and the majority of patients (80.9%) were considered asymptomatic or mild pneumonia (12). The case fatality ratio was 2% (12), 2.3% (8), 3.46% (13) and elderly men with underlying diseases were at a higher risk of death (13). As of the time of the up to date this article, March 29, 2020, COVID-19 pandemic was declared by the World Health Organization (WHO) in more than 200 countries (most prevalent in the United States, Italy, China, Spain, Germany, Iran, France) in worldwide (14). In Iran, the first case of COVID-19 was reported on Day 30 of month 12 (Bahman) in Iranian calendar from Qom, and we used the reported data until March 29, 2020 (15). As of the time of the up to date this article, March 29, 2020, According to the Daily Reports in Iran, 38309 COVID-19, 2640 deaths were reported. in recent weeks, Firstly, universities and schools, then public places and shrines were closed. People are referring to health centers and hospitals, and the public is almost alarmed by the epidemic of panic and inaccurate reporting in cyberspace. The recurring and important questions are: How is the size of the epidemic of COVID-19 in Iran and how long and when will the epidemic go down? We cannot answer these questions with certainty, but it can be investigated in terms of pathogenic behavior (coronavirus), host conditions, behavior (human) and environmental factors of coronavirus transmission, daily reports of definitive COVID-19 patients released by Iran Ministry of Health and Medical Education and the use of modeling given the assumptions and the percentage of error. Indeed, although the models are different, multiple, and changeable in nature and do not insist on the correctness of the forecasts, the decision-making conditions for health policy-makers and authorities are more transparent and helpful (16). This study aimed to model and determine the epidemic trend and predict COVID-19 patients in Iran using mathematical and statistical modeling.

## Materials and methods

This study is a secondary data analysis and mathematical modeling based on a research proposal approved by Shahrekord University of Medical Sciences (Code of Ethics Committee on Biological Research IR.SKUMS.REC 1398.254) (17).

For the statistical analysis of definitive COVID-19 patients in Iran, daily reports of the Ministry of Health and Medical Education were used. The definitive diagnosis of COVID-19 was made using virus isolates from patients’ biological samples in patient with respiratory symptoms and confirmed by the Reference Laboratory located in the School of Public Health, Tehran University Medical of Sciences and Pasteur Institute of Iran were used (18). Patient population growth, epidemic curves, and recovered and deceased individuals were used to conceptual framework of epidemic and predict the COVID-19 epidemic trend. we used classical infectious disease (Susceptible→Exposed→Infected→Removed: SEIR) model in this frame work (19). Different scenarios were designed and implemented for modeling and forecasting. First, based on a search for reliable sources of disease trends and epidemic curves across the world, the curve of Iran was also drawn (10,16,20). Focused and scientific group discussion sessions were held with experts on epidemiology, biostatistics, and mathematics, infectious diseases specialists as well as healthcare managers on the topic, the scenarios were discussed and agreement was reached on the application of the final scenarios. to predict the growth of epidemic different models were used. In the first scenario, the most optimistic estimation and control of the epidemic was during an incubation period (van model) and considering the ideal model. In this scenario traced contacts are isolated immediately on symptom onset (and not before) and isolation prevents all transmission were used. In the second scenario, an intermediate and fit-to-data model (Gompertz) was used. In the third scenario, the use of the growth rate is greater than the first and second models, and in fact opposite to the first scenario (LSE). To select the scenarios, fit the data with the models and growth rate of the cases were used. The Gompertz growth, von Bertalanffy growth equation and curve fitting by LSE method with cubic polynomial for Epidemic forecasts were run in MATLAB software. Models are presented as the following differential equations:

Gompertz Differential Equation:

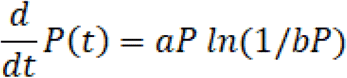

Von Bertalanffy’s differential growth equation:

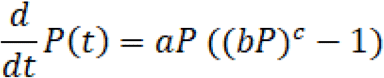

Cubic Polynomial Polynomials:

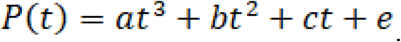

Where P represents the number of individuals in each population, *a, b, c*, and *e* represent unknown parameters and t time. 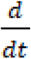 is a derivative of time (20-22).

The unknown parameters are estimated by running the fminsearch, a MATLAB function, which is a least squares algorithm. We perform the estimation of the parameters based on the official reported data of infected, cured and dead cases. The estimated values of the parameters for different scenarios are reported in Tables 2. Also for fitting data and solving the equations we used MATLAB software. MATLAB codes are presented in the supplementary file. The basic reproduction number R_0_ was calculated by the following formula (23):

**Table 2.** Estimated Parameters (values of the inputs used) for modeling

| Parameter<br>Model | infected cases |  |  | recovered cases |  |  | death cases |  |  |
| --- | --- | --- | --- | --- | --- | --- | --- | --- | --- |
| | $a$ | $B$ | $c$ | $a$ | $B$ | $c$ | $a$ | $B$ | $c$ |
| Gompertz | 0.1 | 2.9939e+04 | - | 0.1 | 1.8107e+04 | - | 0.0660 | 2.6710e+04 | - |
| VonBertalanffy | 0.8702 | 2.7109e-05 | 0.2109 | 0.5564 | 7.5133e-05 | 0.3708 | 0.7321 | 2.2840e-05 | 0.1239 |

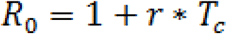

Where, T_c_ and r are the mean generation interval of the infected and the growth rate, respectively and we considered T_c_ = 7.5 and r = 0.1. The growth rate of Gompertz’s model is r=0.1, so the number of R_0_ is 1.75. All estimated are based on current trends, Sampling of severe cases, hospitalization and tip of iceberg spread disease and asymptomatic, mild and moderate cases could not be calculated.

## Results

Frequency of daily statistics of COVID-19 (definite new cases), number of deaths and recovered cases) in Iran are shown in Table 1. The trend of epidemic spread in Iran (daily linear and cumulative trend) are illustrated in Figure 1. According to data released on COVID-19 in Iran as of 10/01/99 (March 29, 2020), the following forecasts until 15/01/99 (April 3, 2020), also until 13 May, 2020 were reported (Figures 2, 3, and 4).

**Table 1:**
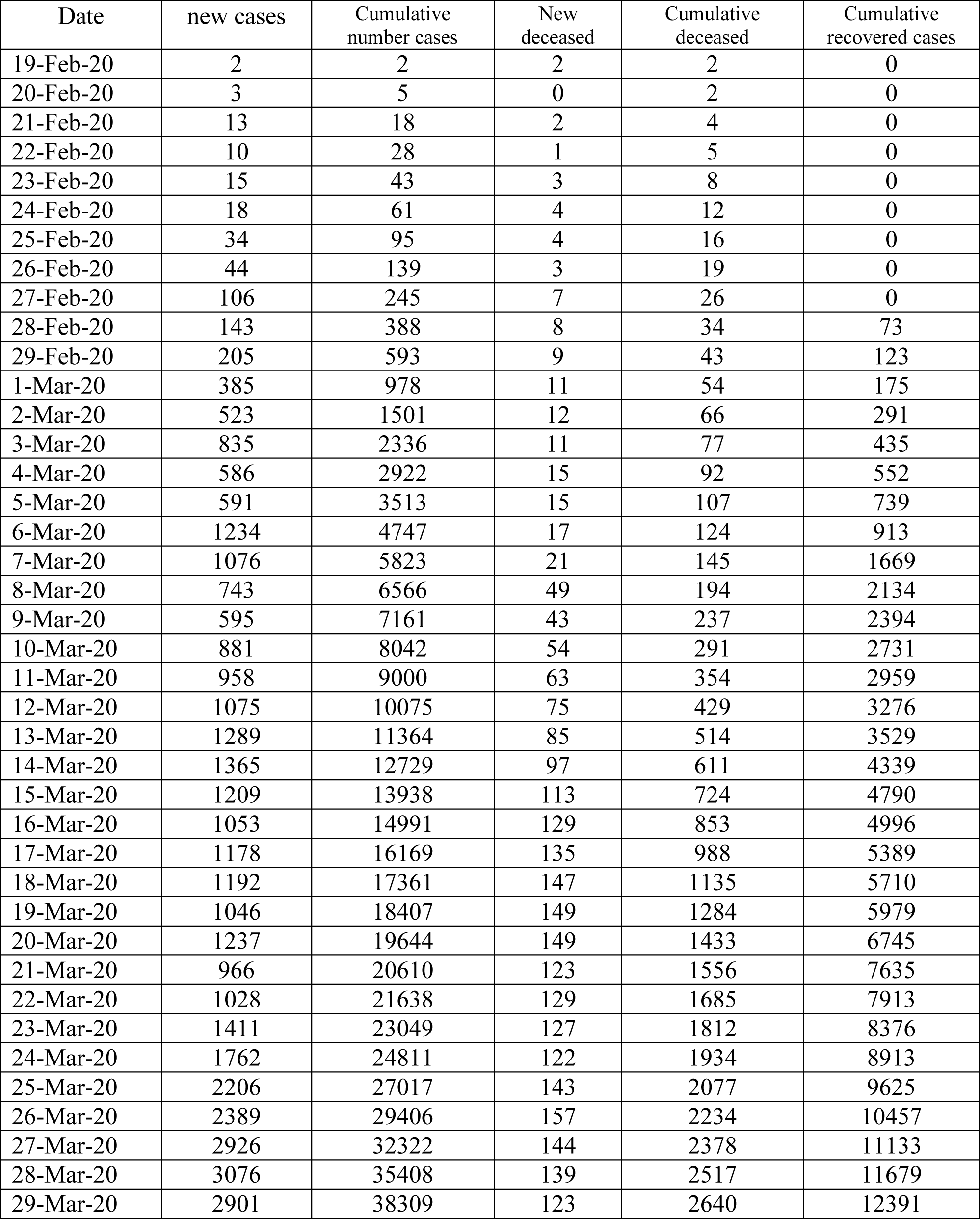
The frequency of COVID-19 new cases, cumulative cases and deceased cases in Iran

| Date | new cases | Cumulative number cases | New deceased | Cumulative deceased | Cumulative recovered cases |
| --- | --- | --- | --- | --- | --- |
| 19-Feb-20 | 2 | 2 | 2 | 2 | 0 |
| 20-Feb-20 | 3 | 5 | 0 | 2 | 0 |
| 21-Feb-20 | 13 | 18 | 2 | 4 | 0 |
| 22-Feb-20 | 10 | 28 | 1 | 5 | 0 |
| 23-Feb-20 | 15 | 43 | 3 | 8 | 0 |
| 24-Feb-20 | 18 | 61 | 4 | 12 | 0 |
| 25-Feb-20 | 34 | 95 | 4 | 16 | 0 |
| 26-Feb-20 | 44 | 139 | 3 | 19 | 0 |
| 27-Feb-20 | 106 | 245 | 7 | 26 | 0 |
| 28-Feb-20 | 143 | 388 | 8 | 34 | 73 |
| 29-Feb-20 | 205 | 593 | 9 | 43 | 123 |
| 1-Mar-20 | 385 | 978 | 11 | 54 | 175 |
| 2-Mar-20 | 523 | 1501 | 12 | 66 | 291 |
| 3-Mar-20 | 835 | 2336 | 11 | 77 | 435 |
| 4-Mar-20 | 586 | 2922 | 15 | 92 | 552 |
| 5-Mar-20 | 591 | 3513 | 15 | 107 | 739 |
| 6-Mar-20 | 1234 | 4747 | 17 | 124 | 913 |
| 7-Mar-20 | 1076 | 5823 | 21 | 145 | 1669 |
| 8-Mar-20 | 743 | 6566 | 49 | 194 | 2134 |
| 9-Mar-20 | 595 | 7161 | 43 | 237 | 2394 |
| 10-Mar-20 | 881 | 8042 | 54 | 291 | 2731 |
| 11-Mar-20 | 958 | 9000 | 63 | 354 | 2959 |
| 12-Mar-20 | 1075 | 10075 | 75 | 429 | 3276 |
| 13-Mar-20 | 1289 | 11364 | 85 | 514 | 3529 |
| 14-Mar-20 | 1365 | 12729 | 97 | 611 | 4339 |
| 15-Mar-20 | 1209 | 13938 | 113 | 724 | 4790 |
| 16-Mar-20 | 1053 | 14991 | 129 | 853 | 4996 |
| 17-Mar-20 | 1178 | 16169 | 135 | 988 | 5389 |
| 18-Mar-20 | 1192 | 17361 | 147 | 1135 | 5710 |
| 19-Mar-20 | 1046 | 18407 | 149 | 1284 | 5979 |
| 20-Mar-20 | 1237 | 19644 | 149 | 1433 | 6745 |
| 21-Mar-20 | 966 | 20610 | 123 | 1556 | 7635 |
| 22-Mar-20 | 1028 | 21638 | 129 | 1685 | 7913 |
| 23-Mar-20 | 1411 | 23049 | 127 | 1812 | 8376 |
| 24-Mar-20 | 1762 | 24811 | 122 | 1934 | 8913 |
| 25-Mar-20 | 2206 | 27017 | 143 | 2077 | 9625 |
| 26-Mar-20 | 2389 | 29406 | 157 | 2234 | 10457 |
| 27-Mar-20 | 2926 | 32322 | 144 | 2378 | 11133 |
| 28-Mar-20 | 3076 | 35408 | 139 | 2517 | 11679 |
| 29-Mar-20 | 2901 | 38309 | 123 | 2640 | 12391 |

**Fig 1.**
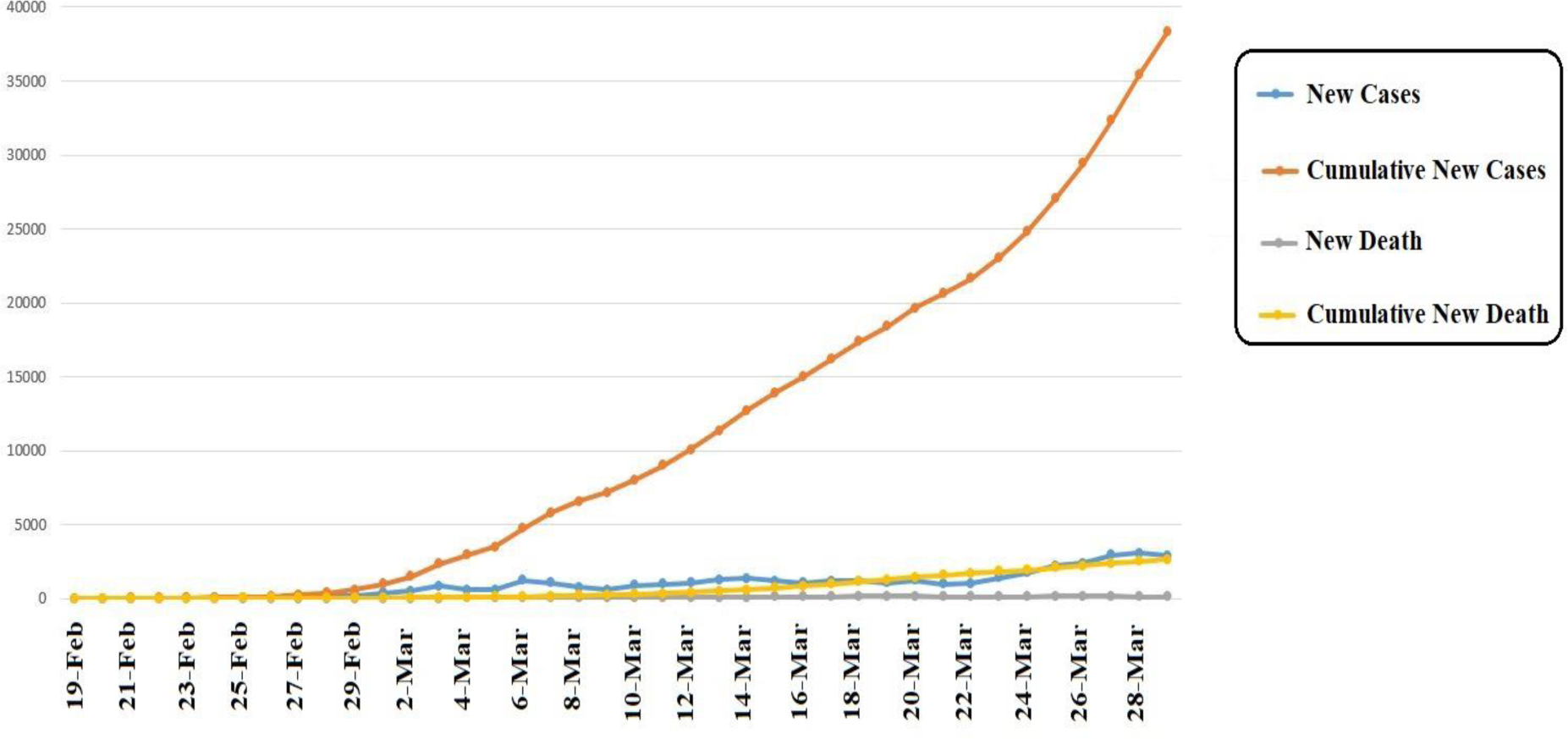
The trend of new and cumulative cases of COVID-19 in Iran.

**Fig 2.**
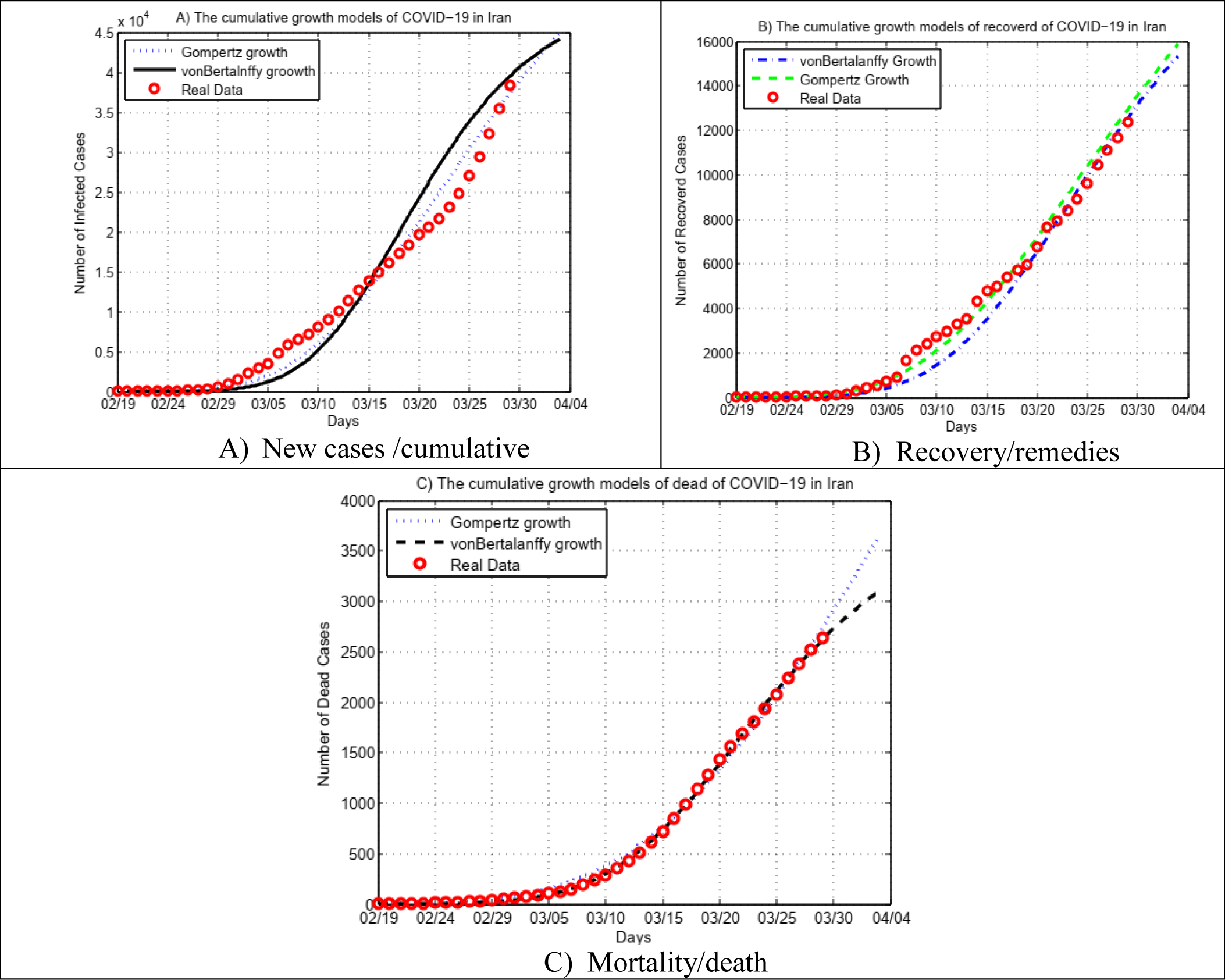
Prediction of COVID-19 by Gompertz and Von Bertalanffy models in Iran.

**Fig 3.**
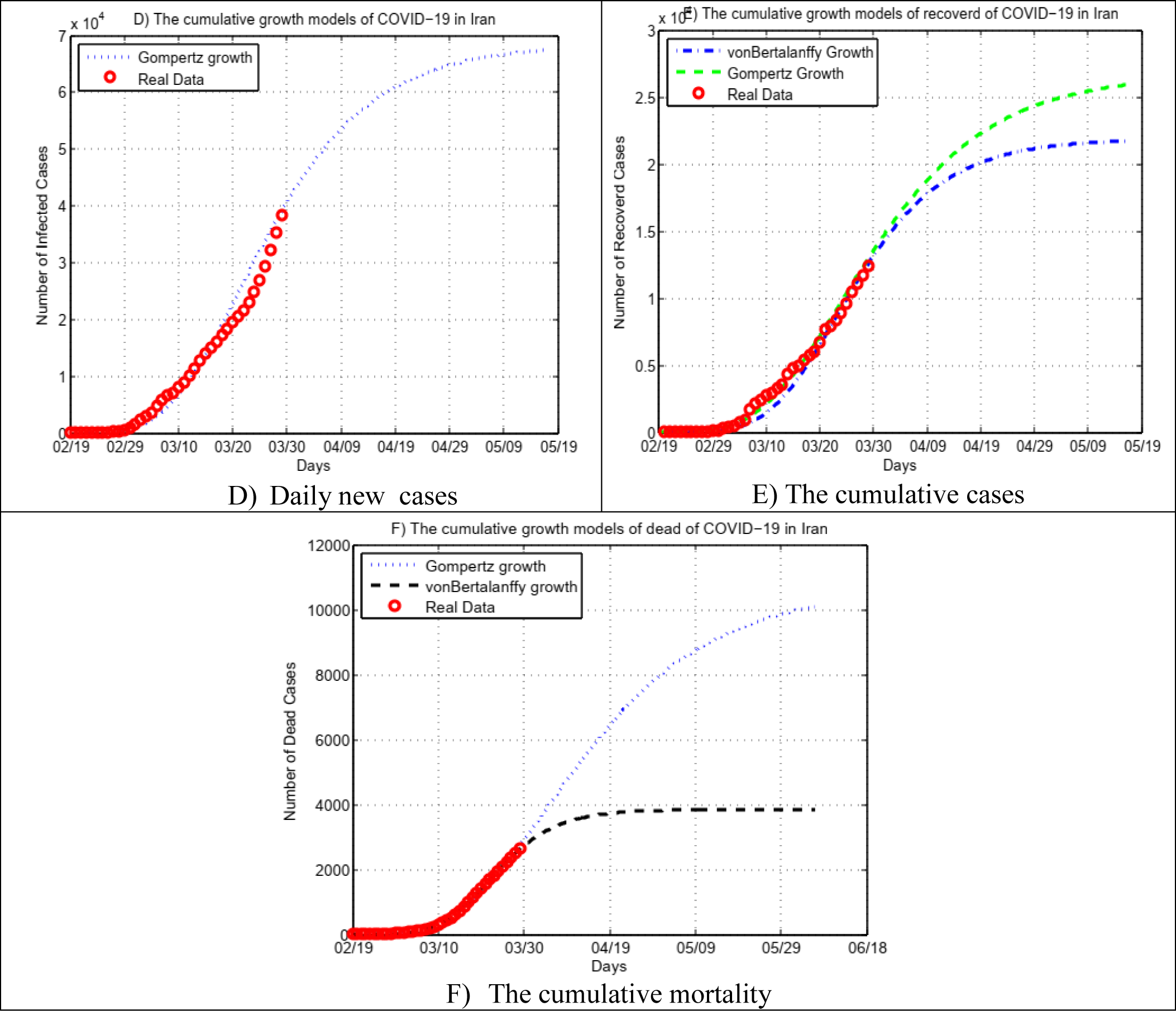
Prediction of the end of COVID-19 epidemic in Iran.

**Fig 4.**
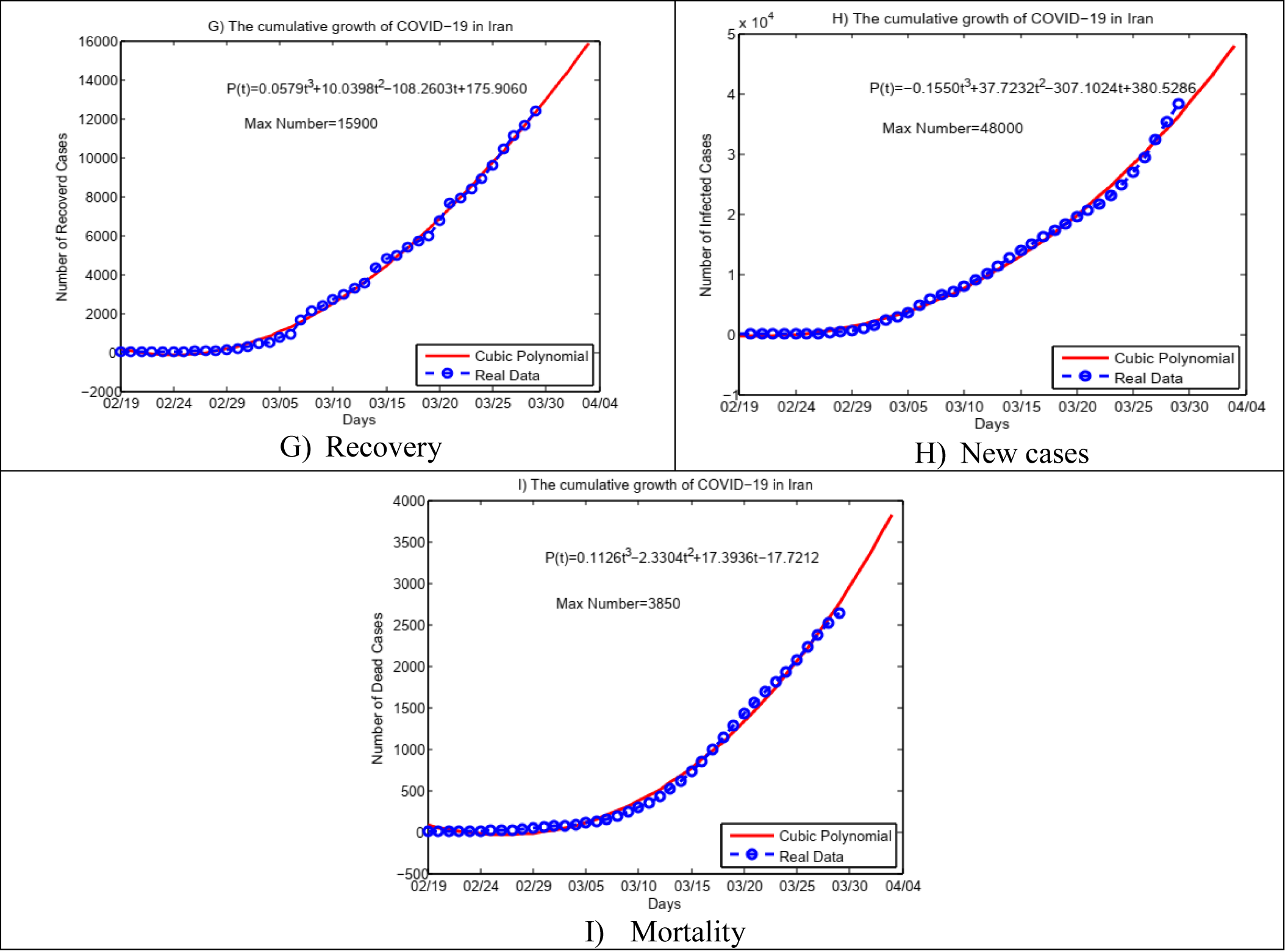
Prediction of COVID-19 by Cubic Polynomial model in Iran.

According to the Gompertz model, in the most optimistic perspective, the maximum number of infected people until April 3, 2020 is 58000 (Figure 2.A). Based on Von Bertalanffy’s growth model (the most ideal model with high isolation of patients and others intervention such as China experience), the maximum patients of COVID-19 are 44200 (Figure 2.A). According to the method of the least squared error, this value was estimated to be 48000 (Figure 4.H).

According to Figure 2.B and the models of Von and Gompertz for prediction of recovered patients, the model estimated the number of recovered individuals to be 15300 and 15950, until April 3, 2020, respectively. Moreover, according to Figure 4.G, the maximum population of recovered individuals was estimated 15900 according to the method of least squared error.

To predict and estimate the number of deaths due to COVID-19, two growth models (Figure 2.C) and the least squared error (Figure 4.I) were used to maximize Von’s growth in the most optimistic manner. According to Von, Gompertz and least squared error method, the number of deaths will be, 3600, 4200 and 4850 individuals on April 3, 2020, respectively.

For prediction end of epidemic, three models of growth were used. Based on Gompertz’s growth model the end of the epidemic is about 13 May, 2020 with the about 87000 patients (Figure 3.D). Based on Von and Gompertz models number of cured cases will be 22000 and 26000, respectively (Figure 3.E). To predict and estimate the number of deaths due to COVID-19 in the end of epidemic Von and Gompertz growth models are used and bsesd on Von model we will have 4900 and based on Gompertz we will have 11000 deaths until 13 May and 1 June, respectively (Figure 3.F).

## Discussion

In this study, we monitor the trend of COVID-19 epidemic, prediction and estimation of the number of patients, R_0_, deaths and recovered individuals were performed and reported based on mathematical and statistical models. Although this prediction may be associated with random errors, it was made with assumptions about the past trends of the COVID-19 epidemic in Iran as well as the behavior of the people and government interventions (Sampling of severe cases and hospitalization). One point that may change our estimates is that we are at the end of the solar year and are less than 10 days longer than the Iranian New Year (Nowruz 1399) and New Year’s with long vacations may increase visits, travels, and other activities. It is a rigorous government and health care intervention such as social distance that may be implemented more and more strictly on these days. Moreover, according to a valid scientific report, delay in the onset of symptoms until the isolation of patients plays an important role in controlling the epidemic. To control the majority of outbreaks, for R0 of 2.5 more than 70% of contacts had to be traced, and for an R0 of 3.5 more than 90% of contacts had to be traced. The delay between symptom onset and isolation had the largest role in determining whether an outbreak was controllable when R0 was 1.5. For R0 values of 2.5 or 3.5, if there were 40 initial cases, contact tracing and isolation were only potentially feasible when less than 1% of transmission occurred before symptom onset (16). Therefore, it is recommended that efforts be made to control the epidemic with greater vigor and urgency and to conduct a daily risk assessment. In the current epidemiological situation in the world and Iran, fear control and avoidance of rumors are very important for the prevention and control of coronavirus.

There are three important points in this epidemic. The first point is to help calm down the atmosphere of society and to avoid tension in societies, and the second is to properly interpret the COVID-19 case fatality ratio (CFR) in Iran and calculate it tactfully. It should be taken into account in interpreting this index, since the denominator of the fraction is only positive cases in hospital beds, and numerator is the number of died positive among the positive cases (of those who are hospitalized and/or die). This index should also be calculated until the end of the epidemic period, and if it is until the end of the epidemic and their outcome (death/recovery) is determined, this indicator will approach the real number. The estimated case fatality ratio among medically attended patients was reported approximately 2% (12) and the true ratio may not be known for some time (25). Compared to this index with Iran, it seems to be much higher in Iran. That requires further investigation and determination of the causes. The third point is to recommend personal hygiene, in particular hand washing and avoiding contact with suspected patients, social distance, discovering unknown cases of infection and early detection, trace direct contact and isolation of patients, which is emphasized by the healthcare system to overcome this disease. One article reported that up to 70% of the supply chain could be cut off and the epidemic could be controlled if contact and isolation, quarantine and isolation were appropriately accomplished (16). The top priorities in Iran are now circular and comprehensive efforts to conduct epidemiological studies and identification of all aspects of the disease (source of disease, reservoir, pathways, infectivity, incubation period, incidence and prevalence, pathogenicity, immunogenicity, herd immunity, causes, epidemic and pandemic pattern, primary and secondary attack rates, response time, time needed for isolation and quarantine, treatment regimens, vaccines and other prevention methods, disease surveillance and statistical reporting) and evidence-based interventions and epidemic control.

Application of experiences in China and South Korea epidemic shows the disease growth in the country to be logistically dispersed and the epidemic will be controlled in the near future, indicating that preventive activities in the two countries have been more useful among all East Asian countries. Obviously, cultural conditions are also effective. Given the prediction and modeling of the number of cases of coronavirus in Iran and because the virus is circulating in the country for at least a few weeks, we will have an ascending trend in the coming weeks. Application of the experiences of China, which took almost 70 days to complete the epidemic curve, has reached a flat level; it is recommended that the following epidemiological recommendations be implemented promptly and that interventions recommended by the World Health Organization be implemented (26).

Up-to-date and accurate data on definitions related to suspected, probable, and definitive people with coronavirus should be collected at all levels of the province’s health care system. Percentage of completion and accuracy of their assessments and data should be monitored precisely and the epidemic curve should be drawn by rural, city, district, province, and other important sectors and be provided to the campus authorities in an updated dashboard format. Data should be carefully recorded and analyzed regarding the pathology, the time of onset of symptoms, natural course of the disease and the outcome of the disease to determine effective strategies to prevent and determine the necessity of intervention to control the spread of the disease at different levels. At all levels of the health care system (governmental and nongovernmental), interventional care, diagnostic and therapeutic interventions, whether compliant or non-compliant, should be conducted for all cases since the beginning of the interventions based on the time of onset of symptoms and type of diagnosis and recorded, and their relationship to outcomes should be analyzed to evaluate the cost-effectiveness of various diagnostic therapeutic approaches. Health care system staff should be empowered to record and train data, especially the use of virtual, remote, and web-based networks. This will certainly improve the quality of data recording. All epidemiological indicators that determine the epidemic pattern, including baseline R zero, attack rate, incubation period, index case, primary cases, secondary cases, and GIS mapping should be determined in provinces, cities, and across the entire country, and epidemic trends should be monitored. We suggested Access to the results of the analysis as well as data should be provided for researchers and experts on the basis of specific protocols available for this purpose in the world and Iran, and a thorough critique and creative theories and ideas should be elicited from all university training and research groups. The models used to predict the end of the epidemic and control it should be evaluated, as well. The results of our study are inconsistent with Zhuang brief report. That study reported the data were collected from the World Health Organization. This report is incorrect and the World Health Organization has not reported it (27). That given the urgency of need for valid and transparent models to inform interventions and policies, it is known that some further considerations like the no consideration or account of systematic cases, testing coverage and time delay to the test results availability, seasonality, and comorbidities have not been include in this study, but they might be feasible to consider them in revisions of the models or future studies. And The progression of disease epidemic across space-time has not been seen in Iran and we used estimation in China to calculated R_0_. We have been no attempt made detection and reporting of cases and deaths in Iran. It could have been performed, but given that the study is already designed, conducted, and being reported, and given the exceptional mortality and morbidity situation, we understand that it would not be feasible now to go back and do something for this issue. However, it can be done possibly for the future similar studies.

## Conclusion

The actual trend of detecting COVID-19 cases in Iran has been increasing and is based on public behavior and government intervention. In this study, estimated are based on current trends, social distance, sampling of severe cases, hospitalization and tip of iceberg spread disease and asymptomatic, mild and moderate cases could not be calculated. Complete reliance on any type of model will lead to systematic and random error, unless modeling provides a prediction with precise and clear assumptions and inputs and outputs. Given the assumptions in Iran, the prediction of the patients on 3 April (Farvardin 15), 2020 with the three growth models of Von Bertalanffy, Gompertz, and the least squared error were estimated at 48200, 52500 and 58000, respectively. The number of deceased COVID-19 patients was also estimated to be 3600 individuals using the Von’s growth model, 4200 ones by Gompertz’’s model and 4850 ones according to the LSE method. If enforcement and public behavior interventions (social distance) continue with current trends, the control and reduction of the COVID-19 epidemic in Iran will be felat from late April 2020, and new cases are expected to decline from mid-April of the following Iranian new year. For prediction end of epidemic, three models of growth were used. Based on Gompertz’s growth model the end of the epidemic is about 13 May, 2020 with the about 87000 patients. Based on Von and Gompertz models number of cured cases will be 22000 and 26000, respectively. To predict and estimate the number of deaths due to COVID-19 in the end of epidemic Von and Gompertz growth models are used and bsesd on Von model we will have 4900 and based on Gompertz we will have 11000 deaths until 13 May and 1 June respectively. This study suggests that government interventions and people’s behaviors determine the persistence of the epidemic and should be addressed with greater responsibility, accountability, rigor, and quality.

## Data Availability

https://mhrc.skums.ac.ir/Index.aspx?page_=form&lang=1&sub=55&tempname=rcmh&PageID=22976&isPopUp=False

http://ethics.research.ac.ir/ProposalCertificateEn.php?id=124057&Print=true&NoPrintHeader=true&NoPrintFooter=true&NoPrintPageBorder=true&LetterPrint=true

## Acknowledgments

We appreciate the Modeling in Health Research Center, School of Health, Deputy of Research and Technology, Shahrekord University of Medical Sciences. Furthermore, we truly appreciate the Ministry of Health and Medical Education, especially the team of daily information reports of COVID-19 in Iran.

## Funding of study

This study was financially supported by the Deputy of Research and Technology of Shahrekord University of Medical Science, Shahrekord, Iran (grant no. 254).

## Conflicts of interest

None.

## Notes

### Competing Interest Statement

The authors have declared no competing interest.

